# Urinary ER Chaperones – Biomarkers of Glomerular ER Stress

**DOI:** 10.64898/2026.09.28.26364164

**Authors:** Andrey V. Cybulsky, Nihad Tousson-Abouelazm, Joan Papillon, Ahsan Alam, Frederic Baroz

## Abstract

Certain glomerulopathies (GN) feature impaired protein homeostasis in the endoplasmic reticulum (ER) of podocytes. Protein misfolding causing ER stress activates the unfolded protein response (UPR), a protective signaling network. Mesencephalic astrocyte-derived neurotrophic factor (MANF) and ERdj3/DNAJB11 are ER chaperones that may be secreted from cells during the UPR. In rats with membranous nephropathy (MN) and focal segmental glomerulosclerosis (FSGS), secretion of MANF or ERdj3 from podocytes into urine identified glomerular ER stress/UPR activation. We address the biomarker potential of MANF and ERdj3 in human MN and FSGS. Glomerular gene expression of MANF and ERdj3 was increased in human MN and FSGS. Urinary MANF and ERdj3 were increased in proteinuric MN and FSGS compared to MN and FSGS in remission (low proteinuria), as well as other GNs, polycystic kidney disease, hypertension and healthy controls. In individuals with proteinuric MN or FSGS, amounts of urinary MANF and ERdj3 correlated weakly with proteinuria; correlation of MANF with ERdj3 was moderate. Males represented ∼60% of persons in each group. eGFR in proteinuric MN and FSGS was >40 ml/min/1.73 m^2^. In MN, there were no significant differences in urinary MANF and ERdj3 between anti-PLA2R positive and negative individuals. In conclusion, proteinuric MN and FSGS show increased glomerular expression of MANF and ERdj3. High urinary levels of these ER chaperones correlate only weakly with proteinuria. In these GNs, urinary ER chaperones are thus non-invasive biomarkers of glomerular/podocyte ER stress. Such biomarkers may eventually be useful in identifying patients for therapies targeting ER protein misfolding.

## Introduction

Glomerulopathies, including membranous nephropathy (MN), focal segmental glomerulosclerosis (FSGS) and diabetic nephropathy feature glomerular visceral epithelial cell or podocyte injury and impaired protein homeostasis (“proteostasis”) in podocytes. In response, podocytes can activate adaptive pathways to enhance their capacity to cope with stress [1-6]. Adaptive responses aim to maintain proteostasis before the onset of maladaptive repair [7,8]. Detection or monitoring of adaptive responses may provide an opportunity for diagnosis of impaired proteostasis, and eventual development of preventive and therapeutic approaches. Podocytes have a complex morphology characterized by cell bodies in the glomerular urinary space, and projecting interdigitating foot processes that are bridged by filtration slit diaphragms at the glomerular basement membrane (GBM). These cells are highly active metabolically and very dependent on proper function of the endoplasmic reticulum (ER), as podocytes produce slit-diaphragm proteins, adhesion molecules and GBM components, all of which are critical to the maintenance of glomerular permselectivity [9,10].

During protein translation, secreted and transmembrane proteins (including the proteins noted above) are translocated into the ER. Here, proteins are covalently modified to attain a correctly folded conformation by the action of chaperones and/or folding enzymes, after which these proteins are transported to the Golgi pathway [8]. Disruption of ER function, due to depletion of ATP or ER calcium, or alterations in the redox environment, leads to accumulation of misfolded proteins, ER stress, and activation of the unfolded protein response (UPR), a compensatory signaling network [1-6]. There are three principal UPR pathways: Activating transcription factor 6 (ATF6) is cleaved by proteases, and cleaved ATF6 enters the nucleus to induce transcription of various genes, including those that encode ER chaperones. Inositol-requiring enzyme 1α (IRE1α) is a protein kinase and RNase that splices X-box binding protein 1 (Xbp1) mRNA to yield a potent transcriptional activator of ER chaperone and other genes. PKR-like ER kinase (PERK) phosphorylates eukaryotic translation initiation factor 2α (eIF2α) to reduce translation and protein load on the damaged ER. The so-called adaptive UPR maintains ER proteostasis, facilitates recovery from stress and may protect against further stresses.

The majority of ER chaperones, such as BiP (GRP78) and GRP94, contain a C-terminal KDEL ER retention motif, which binds to the KDEL receptor. However, a few ER chaperones lack KDEL and are reported to be secreted from cells [2,11]. The roles of secreted chaperones have not been established conclusively, but secreted chaperones may be involved in maintaining extracellular proteostasis [11-13]. The ER HSP40 co-chaperone, ERdj3 (or DNAJB11), was identified as a secreted chaperone that is upregulated as part of the UPR [13]. In the kidney, ERdj3 is important in the maturation of polycystin-1 and uromodulin [11]. ERdj3 can bind to misfolded proteins in the ER and deliver them to BiP to facilitate folding. During ER stress, when the BiP machinery is overwhelmed with misfolded proteins in the ER, ERdj3 remains bound to misfolded client proteins and accompanies them extracellularly to reduce their proteotoxic aggregation [2,13]. Mesencephalic astrocyte-derived neurotrophic factor (MANF or Armet) is another ER chaperone that is upregulated during the UPR and can be secreted [14]. MANF lacks KDEL, but contains a C-terminal RTDL sequence that can potentially bind to KDEL receptors with lower affinity compared with KDEL. Thus, it is believed that that under non-stressed conditions, MANF is retained in the ER and is not secreted, but during ER stress, KDEL chaperones would saturate binding to KDEL receptors, allowing secretion of non-KDEL proteins, including MANF [15]. Both ERdj3 and MANF are widely expressed, including in podocytes and other kidney cells [11,16].

There is evidence for ER stress and activation of the UPR in hereditary and acquired human glomerulonephritis (GN) and in their experimental counterparts [2-6,16,17]. These include primary human MN – passive Heymann nephritis (PHN) in rats, and human FSGS – puromycin aminonucleoside nephrosis (PAN) in rats and adriamycin nephrosis in mice. In human adults, primary MN and FSGS are common causes of nephrotic syndrome, and important causes of chronic kidney disease [18-22]. MN is often caused by anti-phospholipase A2 receptor (PLA2R) antibodies targeting PLA2R in podocytes, but several other target antigens have been identified. Studies in experimental MN and FSGS have demonstrated that activation of the UPR can attenuate podocyte injury and proteinuria [2,23,24]. In human MN, podocytes show a dilated and expanded ER [25] and increased ubiquitin content [26], in keeping with protein misfolding and ER stress. Increased glomerular expression of ER stress markers, such as BiP or C/EBP homologous protein (CHOP), was shown in kidney biopsies of patients with MN and FSGS, as well as other GNs [2,27,28]. Furthermore, human MN, FSGS, IgA nephropathy and diabetic nephropathy showed increased glomerular expression of many ER/UPR and autophagy genes, a significant number of which were shared among these diseases [29].

Given that ER stress may play a potentially important pathogenic role in glomerular disease, non-invasive biomarkers for detecting glomerular ER stress would be advantageous. Indeed, angiogenin, MANF and cysteine-rich with epidermal growth factor-like domain protein-2 (CRELD2) have been proposed as urinary glomerular or tubular ER stress biomarkers in experimental models or human kidney diseases [11,16,17,30-32]. Since podocytes reside in the urinary space, we hypothesized that podocyte injury in GN may be associated with secretion of MANF and ERdj3 from podocytes into the urine, and that ER chaperones in the urine reflect glomerular ER stress. In support of this proposition, in cultured podocytes, ER stress increased ERdj3 and MANF intracellularly and extracellularly. Furthermore, ERdj3 and MANF secretion was blocked by the secretory trafficking inhibitor, brefeldin A [16]. Urinary ERdj3 and MANF increased in rats injected with tunicamycin (in the absence of proteinuria). In experimental MN (PHN) and FSGS (PAN), there were increases in glomerular ER stress, and appearance of ERdj3 and MANF in the urine, coinciding with the onset of proteinuria. Rats with PHN were treated with the chemical chaperone, 4-phenyl butyrate (PBA), starting at the time of disease induction, or after disease was established [16]. In both protocols, 4-PBA, which enhances ER protein folding, significantly reduced proteinuria and urinary ER chaperone secretion. Thus, it was concluded that urinary ERdj3 and MANF reflect glomerular ER stress, and the therapeutic response to 4-PBA could be monitored by urinary ERdj3 and MANF [16].

In the present study, we address the biomarker potential of MANF and ERdj3 in human MN and FSGS. We show that in proteinuric human MN and FSGS there are high urinary levels of ER chaperones, which correlate only weakly with proteinuria. Our results support the view that urinary ER chaperones are biomarkers of glomerular/podocyte ER stress.

## Patients and methods

### Patient urine samples and protein and creatinine measurements

Human urine samples and patient data were collected in nephrology outpatient clinics in hospitals, including the McGill University Health Centre (MUHC), and were provided by the NIDDK Central Repository, a program of the National Institute of Diabetes and Digestive and Kidney Diseases, in the context of a CureGN consortium ancillary study [33]. A few MN samples was also provided by Dr. Laurence Beck (Boston University). The work was approved by the

Research Ethics Board at the MUHC (13-411-BMA). Patients were included if age was between 18 and 75, and estimated glomerular filtration rate (eGFR) was >40 ml/min/1.73m^2^. Collection of samples at the MUHC occurred from 2011 up to 2021. Samples from CureGN were collected prior to 2021. The principal study groups were MN with high urine protein/creatinine ratio (UPCR), and FSGS with high UPCR (see Results). Control groups included MN with low UPCR, FSGS with low UPCR, other GNs, polycystic kidney disease (PKD), primary hypertension (HTN) and healthy individuals. CureGN provided 23-24 samples in each of the MN and FSGS – low and high UPCR groups; other samples (N=97) were procured locally (total N=192). We performed a preliminary study that measured urinary MANF in 9 patients with proteinuric MN, 10 with PKD, 4 with HTN and 5 healthy controls. Based on these preliminary data, we estimated that the adequate sample size would be 11 subjects per group (α=0.05, β=0.2).

All urine samples were stored at -80°C. Urine samples collected in hospital were centrifuged post-collection (1800 x g for 10 min) to remove cell debris, and then stored at -80°C. Urine protein measurements were provided by clinical hospital laboratories or by the CureGN consortium. Urinary creatinine was measured in clinical hospital laboratories; in some samples, we measured creatinine using a kit that employs a picric acid-based method (Cayman Chemical Co.), as described previously [16]. Creatinine concentrations were obtained using the standard curve equation of best fit, and the UPCRs were calculated in g/g.

### Antibodies

Rabbit anti-ERdj3 antibody (15484-1-AP RRID:AB_2094400) was from Proteintech (Rosemont, IL). Rabbit anti-MANF antibody (PAB13301 RRID:AB_10546841) was from Abnova (Walnut, CA). Reactivity of ERdj3- and MANF-specific antibodies with their target proteins was previously validated by showing reductions in signal in shRNA-treated cells [13] or knockout mice [30].

### Immunoblotting

Urine samples were subjected to SDS-PAGE [16]. The samples were standardized to urine creatinine excretion, such that the urine volumes applied to each gel lane reflected the same amount of urine creatinine (usually 5 μg). A urine sample was run on each gel as an internal standard for densitometric quantification (i.e. the same sample was run on every gel). Proteins were electrophoretically transferred to a polyvinylidene fluoride membrane of 0.2 μm pore size. After blocking, membranes were incubated with primary antibody overnight (4°C), followed by horseradish peroxidase-conjugated secondary antibody for 1 h (22°C). Chemiluminescence signals were detected in a ChemiDoc Touch Imaging System (Bio-Rad; Mississauga, ON). Membranes were then incubated with the second primary antibody and secondary antibody, as above, and chemiluminescence signals were detected [16]. A representative immunoblot is shown in Fig. S1.

Density of specific bands was measured using ImageJ software [16]. The densitometry value of the internal standard was set to 1.0 and densitometry values of the other values of the other samples are expressed as a ratio of the standard. We took precautions to perform quantification safeguarding the linearity of signal intensities.

### Dataset analyses

The publicly accessible datasets GSE47183 (“JuCKD-Glom”; Nephroseq) [29,34] and GSE108109 [35] were used for the expression analysis of glomerular MANF and ERdj3 genes. The dataset GSE108112, which is related to GSE108109 [35], was used for the expression analysis of MANF and ERdj3 genes in the tubulointerstitium. These datasets contain microarray gene expression data (mRNAs) of laser-captured glomeruli or tubules from human kidney biopsies [29]. The data are presented as the fold-increase of gene expression in disease above healthy control. It should be noted that since these datasets have been generated/published independently, we did not pool the data together and we report the datasets separately. The P-values, which we present in this manuscript, are those calculated and reported within the datasets by their original authors [29].

### Statistics

Experimental data that we generated are presented as mean ± standard error (SE). Comparisons between two groups were done by a two-tailed Student’s t-test. For three or more groups, statistical differences were assessed using one-way analysis of variance. Where significant differences were found, post-hoc analyses were performed using Sidak’s multiple comparisons test.

To evaluate the diagnostic discrimination power of urinary MANF and ERdj3, analyses were performed comparing patients with proteinuric MN and proteinuric FSGS (high UPCR) to all control cohorts. To ensure clinical specificity, analyses were conducted in distinct parallel cohorts, including proteinuric MN versus controls, and proteinuric FSGS versus controls. To assess the discriminative performance of continuous biomarker levels, the area under the receiver operating characteristic (AUROC) curve was calculated. Since our specific datasets exhibited class imbalance, the area under the precision recall curve (AUPRC) was also utilized. Given the class imbalance in our datasets and the proof-of-concept nature of the study assuming equal importance to false positives and false negatives, the optimal diagnostic threshold for biomarker levels was determined by maximizing the F1 score, representing the harmonic mean of precision (positive predictive value) and recall (sensitivity). At this optimal threshold, comprehensive diagnostic metrics were computed and reported, including recall, precision, specificity (true negative rate) and negative predictive value. To quantify the statistical uncertainty of the discriminative metrics, 95% confidence intervals for both the AUROC and AUPRC point estimates were generated using a non-parametric bootstrapping approach of 1,000 iterations. All statistical analyses were performed using Python (v3.11, Python Software Foundation), utilizing the scikit-learn library (v1.6) for diagnostic metric computation and bootstrapping procedures.

## Results

### Glomerular MANF and ERdj3 gene expression is increased in human glomerulopathies

Previously, we analyzed several human gene expression datasets and demonstrated upregulation of multiple ER and UPR genes in MN, FSGS, IgA nephropathy and diabetic nephropathy [24,29,36]. To set the stage for measurement of the urinary ER chaperones, MANF and ERdj3, we analyzed gene expression of MANF and ERdj3 in MN and FSGS using the GSE47183 (JuCKD-Glom) [34] and GSE108109 datasets [35]. Both datasets contain microarray gene expression data of laser-captured glomeruli from human kidney biopsies, and provide the fold-change in gene expression in MN and FSGS groups compared with healthy controls (Table 1). In GSE47183 and GSE108109, MANF gene expression was increased significantly in both MN and FSGS (Table 1). ERdj3 was not examined in GSE47183, but its expression was increased significantly in both MN and FSGS in GSE108109 (Table 1).

**Table 1.**
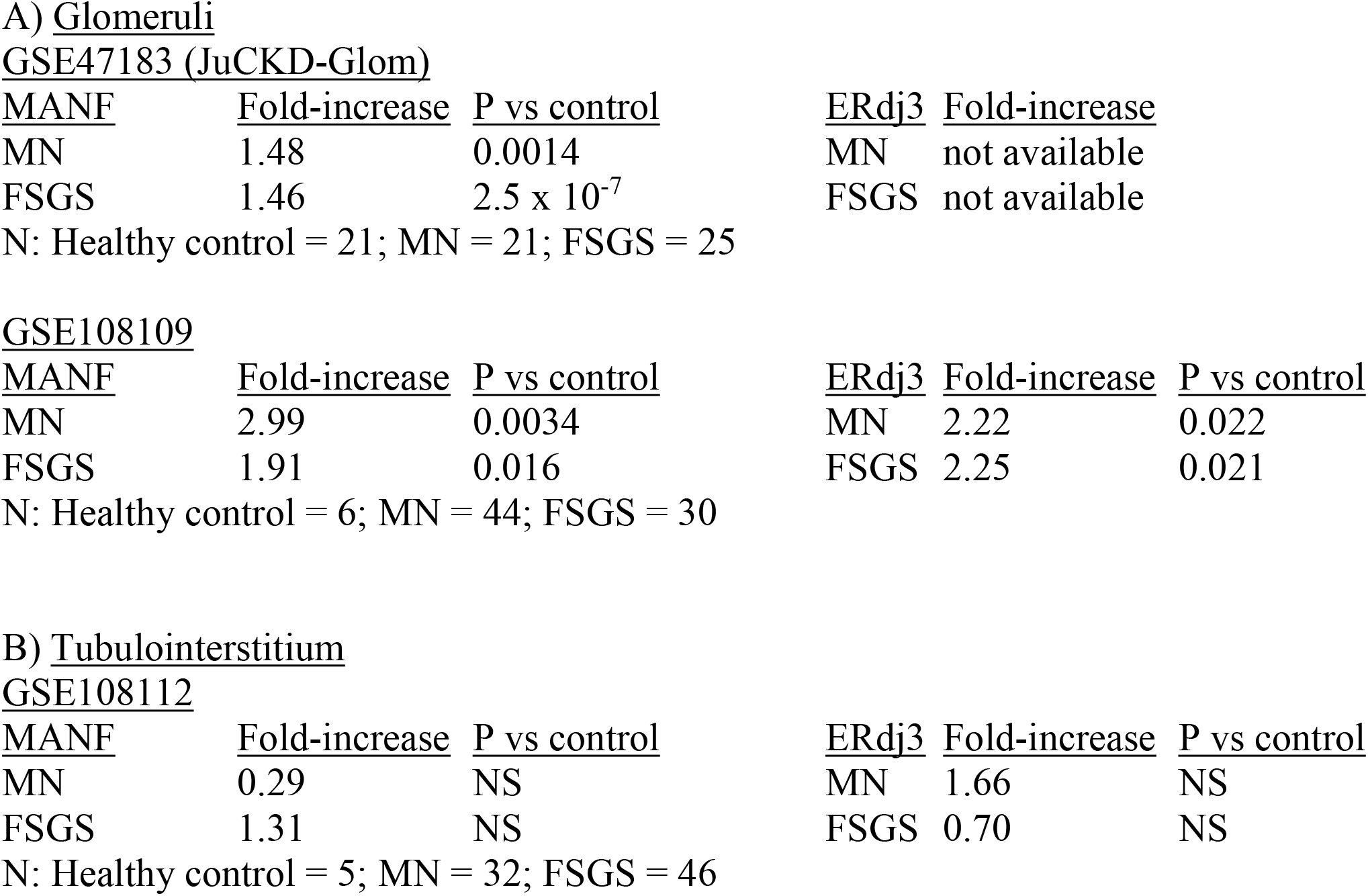
Expression of MANF and ERdj3 mRNAs in human kidneys. Results are based on analyses of GSE47183 and GSE108109 (glomerular datasets), as well as GSE108112 (tubulointerstitial dataset). Values are presented as fold-increase of expression above healthy control in MN or FSGS. NS, not significant.

In GSE47183, the mean age of patients with glomerular disease was ∼50 years, mean eGFR was ∼80 ml/min/1.73 m^2^ and 46% were female. In healthy controls, the values were 47 years, 105 ml/min/1.73 m^2^ and 43%, respectively. In GSE108109, the mean age of patients with glomerular disease was ∼47 years, mean eGFR was ∼77 ml/min/1.73 m^2^ and 33% were female. In healthy controls, the values were 63 years, 108 ml/min/1.73 m^2^ and 57%, respectively [29].

Next, we asked if the glomerular pathophysiological changes in GN could secondarily affect tubulointerstitial expression of ER genes, specifically MANF and ERdj3. The dataset GSE108112 is related to GSE108109, and it contains data on tubulointerstitial gene expression in MN and FSGS based on laser-captured tubular segments from human kidney biopsies [35]. In contrast to changes in glomerular MANF and ERdj3 gene expression, analysis of GSE108112 showed no significant increases in tubulointerstitial gene expression of MANF and ERdj3 in MN and FSGS (Table 1).

### Urinary MANF and ERdj3 are increased in proteinuric glomerulopathies

ER chaperones were measured in urine samples of patients with proteinuric MN and FSGS or “high UPCR” (range 2-17 g/g), which is typically associated with active disease. These measurements were compared with values obtained in several control groups, including MN and FSGS in remission (“low UPCR”; <0.3 g/g), other GNs, PKD, HTN and healthy controls. The other GN group includes IgA nephropathy, minimal change disease, post-infectious GN, C3GN, secondary MN and Alport nephritis. Urinary MANF and urinary ERdj3, as well as UPCR were all significantly greater in the proteinuric MN and FSGS groups, compared with all control groups (Fig. 1).

**Figure 1.**
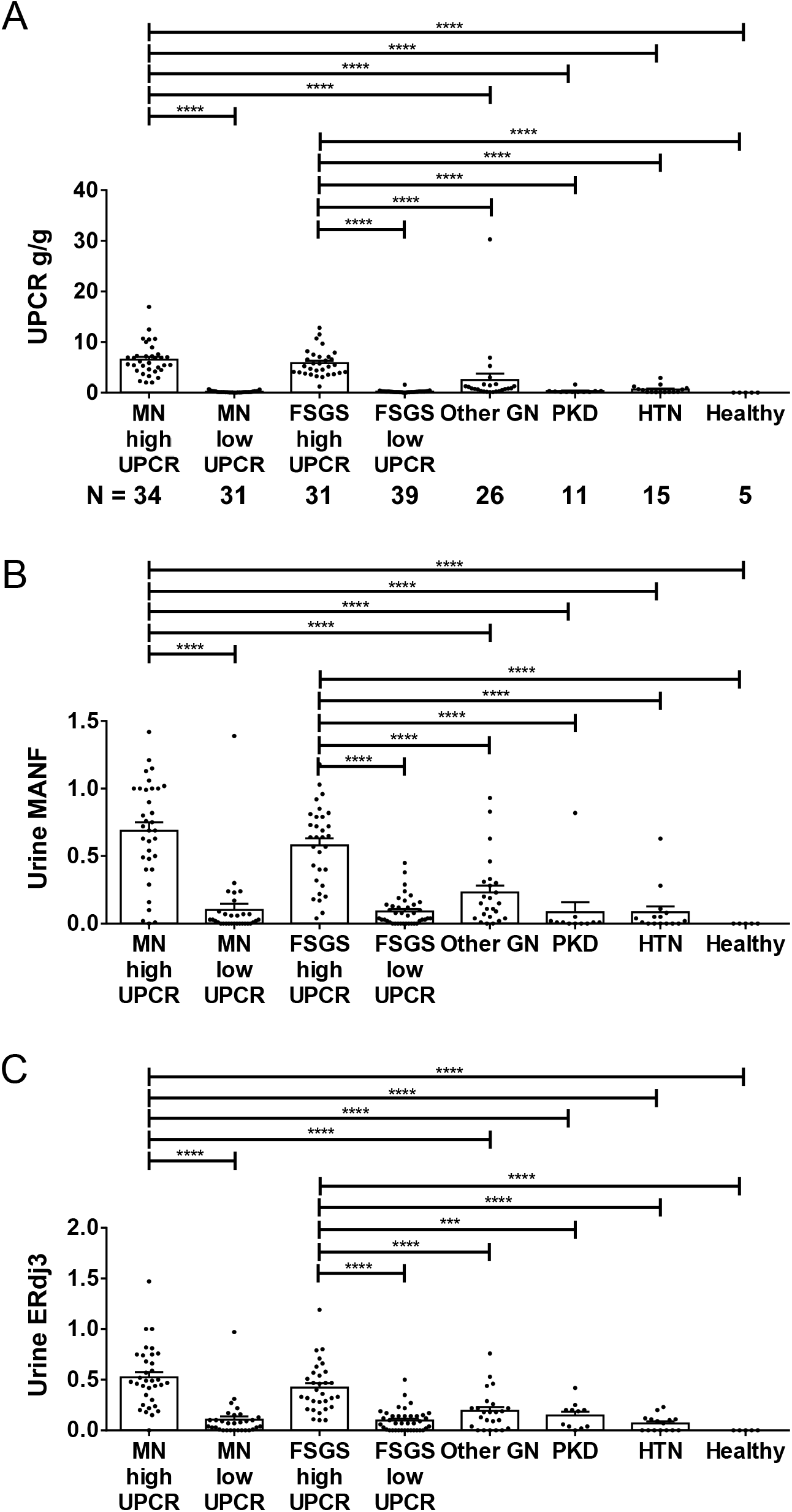
Urinary MANF and ERdj3 are increased in proteinuric MN and FSGS. A) UPCR values in the groups. Subjects with primary MN and FSGS were divided into groups of proteinuric disease (high UPCR) and disease in remission (low UPCR). The other GN group includes IgA nephropathy, minimal change disease, post-infectious GN, C3GN, secondary MN and Alport nephritis. The number of subjects in each group (N) is given below each column. B) Urinary MANF (arbitrary units). C) Urinary ERdj3 (arbitrary units). ****P<0001.

Next, we examined if in the individual patients with proteinuric MN and FSGS, the levels of urinary MANF and urinary ERdj3 correlated with UPCR. Interestingly, analyses showed that these correlations were relatively weak for all groups (Fig. 2). In contrast, urinary MANF correlated moderately with urinary ERdj3 in both proteinuric MN and FSGS groups (Fig. 3).

**Figure 2.**
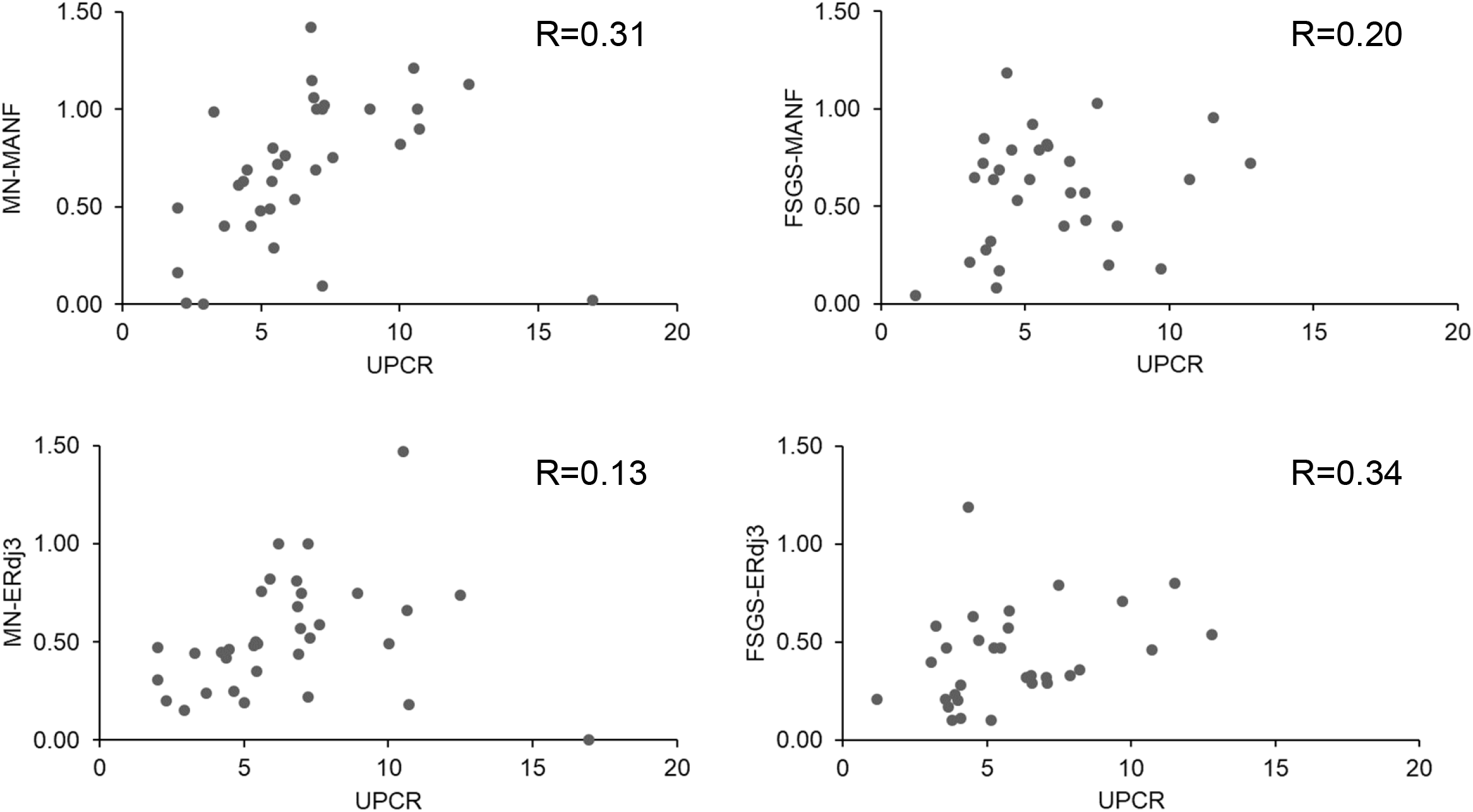
Correlation of urinary MANF and ERdj3 (arbitrary units) with UPCR (g/g) in the MN high UPCR and FSGS high UPCR groups. The correlations are relatively weak.

**Figure 3.**
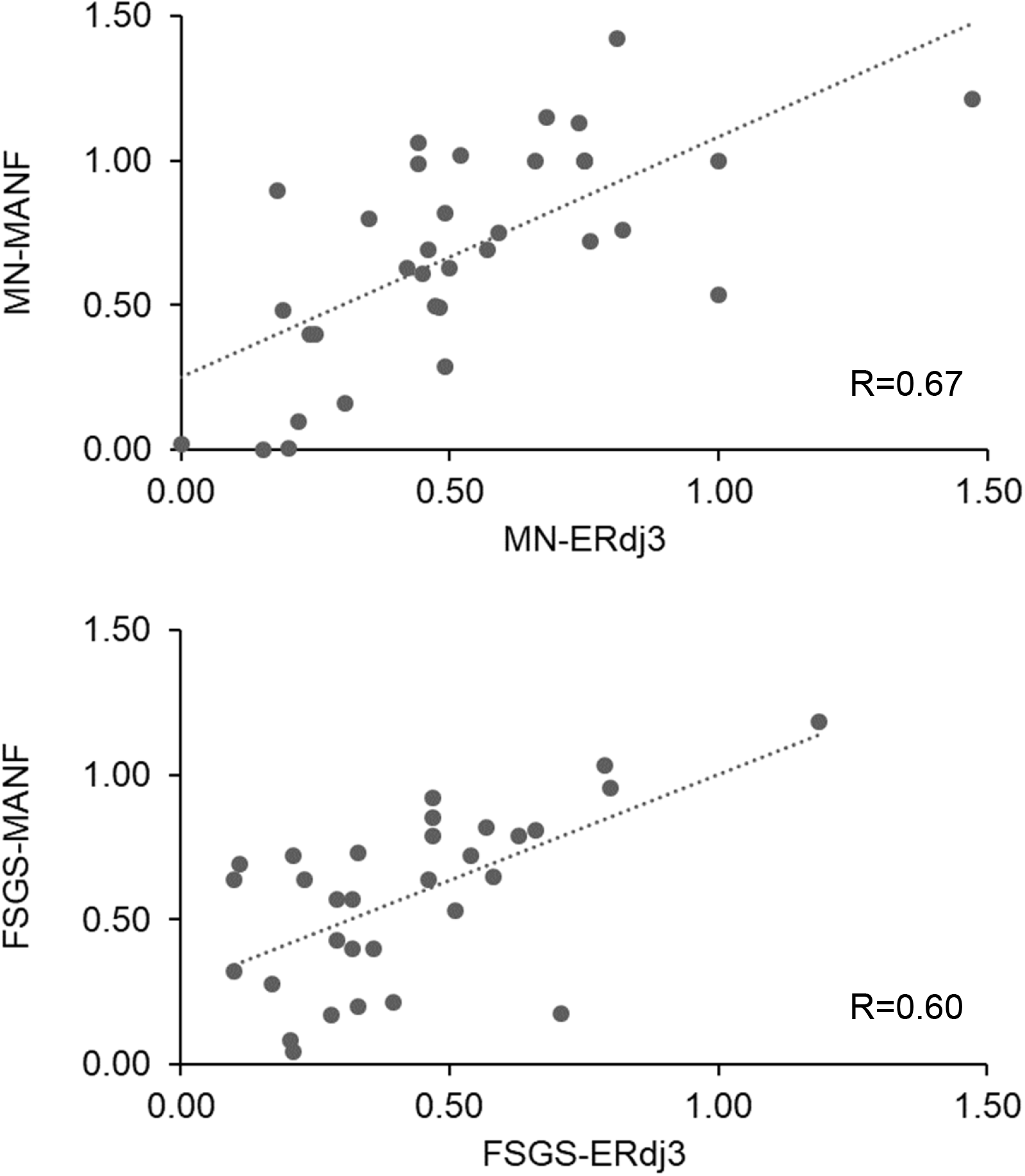
Correlation of urinary MANF with ERdj3 (arbitrary units) in the MN high UPCR and FSGS high UPCR groups. The correlations are moderate.

ROC curves were generated for urinary MANF and ERdj3 in proteinuric MN and FSGS, based on the data in Fig. 1. In both glomerulopathies, binary classification demonstrated excellent discriminatory power, with AUROCs ranging from 0.89 to 0.91 (Fig. 4). To account for the dataset imbalance (∼2.5-3.0 negative cases for every 1 active case), the AUPRC was also evaluated, demonstrating robust performance ranging from 0.74 to 0.79. For proteinuric MN, the optimal F1 scores were 0.81 for MANF and 0.78 for ERdj3, achieved at thresholds of 0.40 and 0.30 units, respectively. At this threshold, MANF demonstrated 82.4% sensitivity, 91.9% specificity, and a positive predictive value of 80.0%. Similarly, ERdj3 achieved 76.5% sensitivity, 91.9% specificity, and a positive predictive value of 78.8%. For proteinuric FSGS, optimal F1 scores reached 0.75 for MANF (threshold 0.40) and 0.74 for ERdj3 (threshold 0.28). At these thresholds, MANF yielded 74.2% sensitivity, 92.6% specificity, and a positive predictive value of 76.7%, while ERdj3 showed 74.2% sensitivity, 91.5% specificity, and a positive predictive value of 74.2%. Consistent with these findings, subjects with positive MANF tests would constitute 82-85% of proteinuric MN and 76-79% of proteinuric FSGS, while positive ERdj3 tests would constitute 74-77% of MN and 61-71% of FSGS.

**Figure 4.**
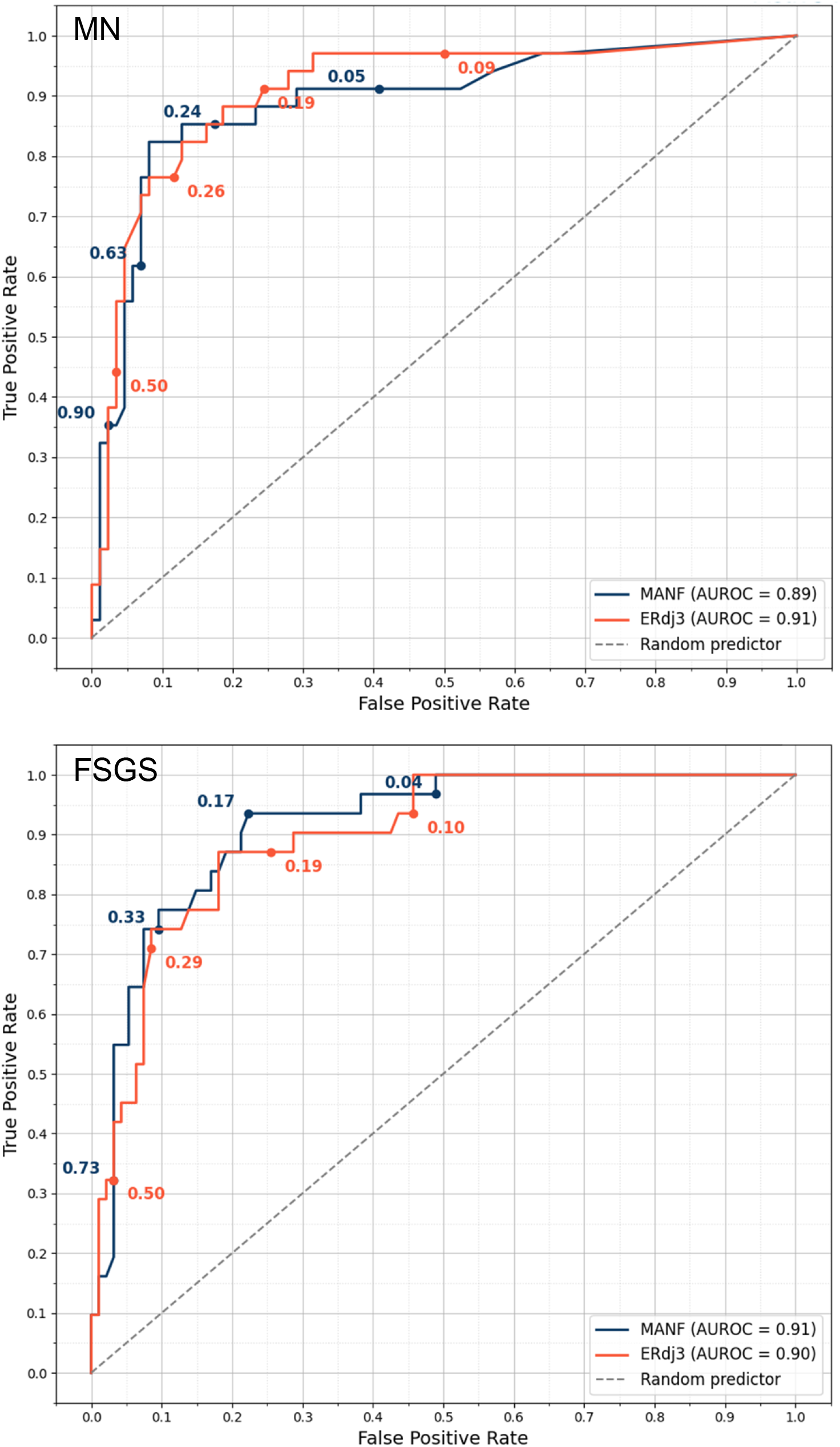
ROC curves for urinary MANF and ERdj3 in proteinuric MN and FSGS, Proteinuric MN was analyzed versus all other groups excluding proteinuric FSGS, and proteinuric FSGS was analyzed versus all other groups excluding proteinuric MN (see Fig. 1). The numbers of subjects in the groups were: proteinuric MN – 34 (28%) vs other groups – 86 (72%), and proteinuric FSGS – 31 (25%) vs other groups – 94 (75%).

The range of UPCR in the other GN group was 0.1-30.0 g/g. There were 4 patients in this group with UPCR >3 g/g, including one with post-infectious GN, one with secondary MN and two with minimal change disease. These patients had values of urinary MANF and ERdj3 >0.3 units. Probably, these patients had significant podocyte injury associated with ER stress, which would account for the higher levels of urinary ER chaperones; thus, these patients may actually be true positives.

### Urinary MANF and ERdj3 do not correlate with anti-PLA2R antibodies

In patients with MN and circulating anti-PLA2R antibodies, it is believed that presence of antibody reflects immunologic disease activity, and antibody titers may be useful in prescribing and following immunosuppressive treatments [18,19]. Measurements of anti-PLAR antibodies were available in a subset of 23 patients with MN. These measurements were presented as positive or negative, and data on antibody titers were not available. Analysis of these patients showed that there were no significant differences in urinary MANF and ERdj3 between anti-PLA2R positive and negative individuals (Fig. 5). UPCR tended to be greater in positive compared to negative subjects, but the difference was not statistically significant (Fig. 5). Thus, urinary ER chaperones and anti-PLA2R antibodies appear to reflect distinct parameters of injury/activity in MN.

**Figure 5.**
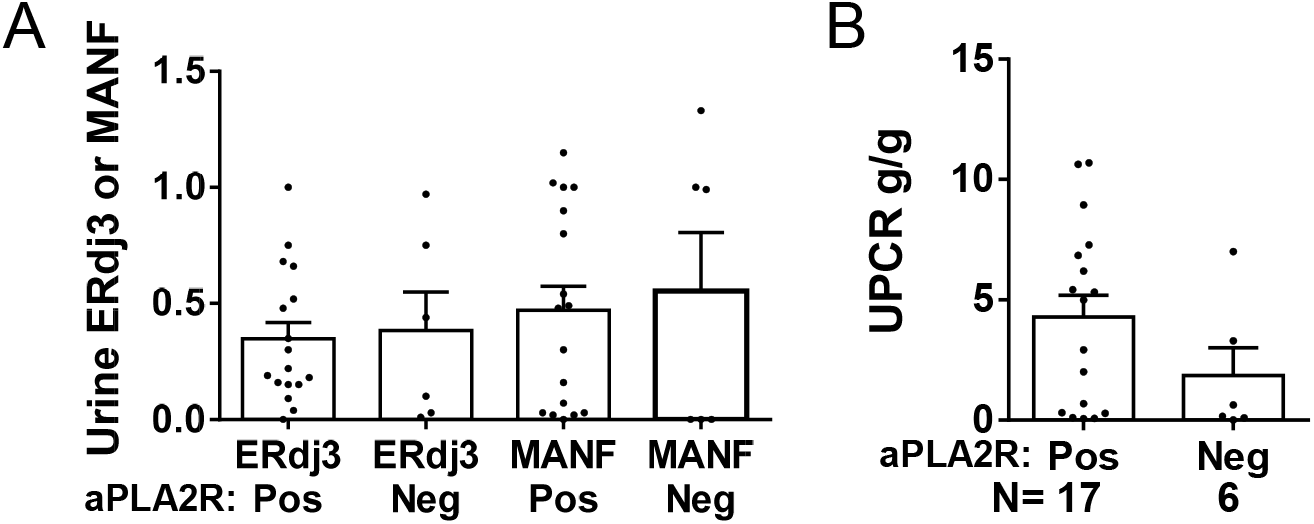
Urinary MANF and ERdj3 do not correlate with anti-PLA2R antibodies in MN (N=23). A)Urinary MANF and ERdj3 (arbitrary units) in anti-PLA2R antibody (aPLA2R) positive (Pos) and negative (Neg) patient groups. B) UPCR. There are no significant differences in urinary ER chaperones and UPCR between anti-PLA2R antibody positive and negative groups.

### Additional analyses

There were minor variabilities in ages of subjects among the various study groups. Subjects with HTN were older compared to proteinuric MN and proteinuric FSGS, while patients in the other GN group were younger than proteinuric MN (Fig. S2). Correlations of urinary MANF, urinary ERdj3 and UPCR with age in proteinuric MN and proteinuric FSGS groups were weak (Fig. S3). Female subjects represented 39.4% of all patients, and there were no significant differences in sex distribution among the groups (Table S1). We also analyzed UPCR, urinary MANF and urinary ERdj3 stratified by sex in proteinuric MN and FSGS (Table S2). There are no significant differences between males and females in all of these parameters, although in FSGS, UPCR tended to be higher in females, while MANF and ERdj3 tended to be higher in males. In summary, age and sex appeared to have minimal effects on the results.

## Discussion

ER dysfunction and impaired proteostasis are associated with the pathogenesis of glomerular diseases; however, non-invasive biomarkers to detect and monitor ER stress are lacking. Our previous studies in experimental MN and FSGS showed that KDEL-deficient ER chaperones, MANF and ERdj3, are candidate urinary biomarkers of ER stress [16]. Specifically, we showed that during ER stress, MANF and ERdj3 were secreted from cultured podocytes through the canonical ER-Golgi pathway, consistent with observations in other cell types [13,37,38]. Since podocytes reside in the urinary space, we asked if the secretion of MANF and ERdj3 from podocytes into the urine can serve as a biomarker of podocyte ER stress in experimental GN, and we demonstrated that levels of these ER chaperones were increased in glomeruli and in urine in the context of podocyte ER stress and proteinuria [16]. Furthermore, in experimental MN, the chemical chaperone, 4-PBA, reduced proteinuria and urinary ERdj3 and MANF [16]. Chemical chaperones provide a more favorable environment in which a protein can fold, by increasing the strength of intramolecular hydrophobic bonds that are buried within a protein or lowering a protein’s free energy state [39]. This may facilitate proper folding and trafficking of membrane and secreted proteins, and prevent the proteotoxic effects of misfolded proteins. In the present study, we show that urinary MANF and ERdj3 are potential biomarkers of glomerular ER stress in primary human MN and FSGS, both of which involve podocyte injury. Glomerular expression of MANF and ERdj3 mRNAs was increased in human MN and FSGS. Levels of urinary MANF and ERdj3 were increased in proteinuric MN and FSGS, compared to MN and FSGS in remission, as well as other GNs, PKD, HTN, and healthy controls. AUROC curves showed that the binary classification model had excellent discriminatory power.

It is well-accepted that proteinuria in GN reflects glomerular injury, often podocyte injury [10,40]. In the present study, subjects with elevated urinary MANF and ERdj3 were among those with substantial proteinuria. However, in the individual subjects with proteinuric MN and FSGS, urinary MANF and ERdj3 levels correlated only weakly with proteinuria. By analogy, in our previous study, urinary MANF and ERdj3 correlated weakly with proteinuria in experimental GN [16]. Thus, it is reasonable to conclude that the presence of urinary MANF and ERdj3 reflects podocyte ER stress, whereas levels of proteinuria may reflect acute or chronic podocyte injury (e.g. disruption of the cytoskeleton or metabolic pathways), as well as pathologic changes to GBM architecture, or altered glomerular hemodynamics [40].

In proteinuric MN, there were no significant differences in urinary MANF and ERdj3 between anti-PLA2R antibody positive and negative individuals. Levels of these urinary ER chaperones covered broad ranges in both groups. The presence of anti-PLA2R antibody is used as a diagnostic marker of MN and titers may be used to follow immunologic activity [18-20]. Our results imply that in MN, glomerular ER stress, as measured by urinary MANF and ERdj3, is separate from immunologic activity. Finally, it should be noted that in humans with proteinuric MN and FSGS, there was moderate correlation between urinary MANF and ERdj3 excretion, although earlier, we found that in experimental MN there was relatively weak correlation between urinary MANF and ERdj3 [16]. These results suggest that in order to establish a diagnosis of glomerular ER stress, it would be preferable to measure both urinary ER stress biomarkers.

A question that arises is whether in MN and FSGS, urinary ERdj3 and MANF were secreted exclusively from podocytes. The datasets that we examined provide gene expression data in glomeruli; thus, changes in gene expression may have occurred in a subset or in all glomerular cells. Nevertheless, since podocytes are the primary sites of injury in MN and FSGS, the gene expression changes most likely reflect changes in podocytes. In GN, tubular ER stress could potentially arise secondary to proteinuria. However, there were no significant changes in tubulointerstitial MANF or ERdj3 gene expression in a human MN and FSGS dataset. In keeping with this result, we showed previously that there were only minimal or no significant differences between expression of MANF and ERdj3 proteins in kidney tubules between rats with MN and controls [16]. In addition, comparable levels of MANF and ERdj3 were excreted in the urine of rats with experimental MN and FSGS, even though MN rats had ∼15-fold greater urinary protein excretion [16]. As stated above, there was only weak correlation between the amounts of urinary ER chaperone excretion and levels of proteinuria in humans, as well as in experimental GN models [16]. This result would also argue against proteinuria being a major stimulus for secretion of chaperones from tubular cells into the urine. In our experience, intrinsic tubular disease may lead to tubular ER stress, but did not result in an increase in urinary ER chaperones. For example, mice with cystinosis, a tubulopathy associated with ER stress due to lysosomal accumulation of cysteine crystals [41,42] did not show an increase in urinary ERdj3 [16]. Mice with experimental PKD demonstrated tubular damage and ER stress [17]; however, in the present study, humans with PKD did not show increases in urinary MANF or ERdj3.

Other studies have addressed ER chaperones as potential ER stress biomarkers. Urinary MANF and CRELD2 (another KDEL-free chaperone) were increased in a mouse model of nephrotic syndrome in which podocyte ER stress was activated by transgenic expression of a C321R laminin mutant in podocytes [30,31]. ERP57 (another KDEL-free chaperone) was detected in the urine of mice with renal fibrosis, as well as patients with diabetic nephropathy and microalbuminuria [43]. Possibly, the interaction of ERP57 with specific secretory client proteins (i.e. extracellular matrix components) may lead to co-secretion of the chaperone. Urinary CRELD2 levels were significantly increased in humans with autosomal dominant tubulointerstitial kidney disease compared with controls [11]. Thus, CRELD2 was proposed as a urinary ER stress biomarker that may be useful in monitoring disease progression in these individuals.

Treatment of human MN and FSGS has relied on relatively non-specific immunosuppressive agents, which target various immune pathways believed to be responsible for triggering disease, but are also associated with significant toxicity [18-22]. Blockade of pathways downstream of immune triggering events, such as the renin-angiotensin system, is effective in reducing progression of glomerular injury; however, additional therapeutic approaches to reduce injury are desirable. Measurement of proteinuria alone does not inform treatment choices [40]. The recognition of protein misfolding and ER stress in various human glomerular diseases, and delineation of this in human patients with non-invasive biomarkers will open an opportunity for testing of pharmacological agents to modulate ER stress pathways, including the UPR, or improve protein folding. Chemical chaperone drugs have been employed with some efficacy in human kidney and non-kidney diseases, and have shown a good safety profile, although chemical chaperones tend to be drugs of low affinity [2,6]. There are also other candidate drugs that target the ER [1,2,8,44,45] and these could potentially be adopted for slowing the progression of MN, FSGS and other glomerular diseases in humans. Our “proof of concept” study indicates that urinary biomarkers of ER stress may eventually be useful in identifying patients for therapies targeting protein misfolding. Future studies will also be required to develop a clinical bioassay for urinary ER chaperones, and address if urinary biomarkers of ER stress predict disease progression.

## Data Availability

All data produced in the present work are contained in the manuscript.

## ACKNOWLEDGMENTS

This work was supported by Research Grants from the Canadian Institutes of Health Research (PJ9-166216 and PJ9-169678) and the Kidney Foundation of Canada, and the Catherine McLaughlin Hakim Chair (AVC).

## AUTHORS CONTRIBUTIONS

AVC and NTA conceived and designed the research. JP and NTA performed experimental work. AA collected and provided urine samples. FB performed statistical analyses. All authors examined the data and results of experiments. AVC wrote the manuscript. All authors reviewed, edited and approved the manuscript.

