## Supplemental Materials for "Urinary ER Chaperones – Biomarkers of Glomerular ER Stress"

**SUPPLEMENTARY MATERIALS**

**Table S1.** Sex distribution among groups.

| | MN high UPCR | MN Low UPCR | FSGS high UPCR | FSGS Low UPCR | Other GN | PKD | HTN | Healthy |
|---|---|---|---|---|---|---|---|---|
| % female | 38.2 | 41.9 | 48.4 | 35.9 | 41.2 | 36.4 | 33.3 | 40.0 |

**Table S2.** UPCR, urinary MANF and urinary ERdj3 (arbitrary units) stratified by sex in proteinuric MN and FSGS. There are no significant differences between males and females in all parameters.

| MN | UPCR, g/g | MANF | ERdj3 |
|---|---|---|---|
| Females | | | |
| Mean | 6.3 | 0.71 | 0.50 |
| SE | 0.5 | 0.08 | 0.06 |
| N=13 | | | |
| Males | | | |
| Mean | 6.7 | 0.67 | 0.54 |
| SE | 0.8 | 0.09 | 0.07 |
| N=21 | | | |

| FSGS | UPCR, g/g | MANF | ERdj3 |
|---|---|---|---|
| Females | | | |
| Mean | 6.1 | 0.48 | 0.36 |
| SE | 0.7 | 0.07 | 0.05 |
| N=15 | | | |
| Males | | | |
| Mean | 5.6 | 0.68 | 0.48 |
| SE | 0.7 | 0.07 | 0.07 |
| N=16 | | | |

**Supplementary Figure Legends**

**Figure S1.** Representative immunoblot showing MANF and ERdj3 in 13 urine samples. Membranes were blotted with anti-MANF antibody and then with anti-ERdj3 antibody. See Methods. Lanes: 1) MN high UPCR, 2) MN high UPCR, 3) MN low UPCR, 4) MN high UPCR, 5) MN high UPCR, 6) FSGS high UPCR, 7) FSGS high UPCR, 8) FSGS low UPCR, 9) MN high UPCR, 10) FSGS low UPCR, 11) FSGS high UPCR, 12) FSGS low UPCR, 13) FSGS low UPCR.

**Figure S2.** Ages of subjects in the experimental groups (years). N values are as in Fig. 1. ****P<0.0001.

**Figure S3.** Correlation of urinary MANF, urinary ERdj3 (arbitrary units) and UPCR (g/g) with age (years) in the MN high UPCR and FSGS high UPCR groups. The correlations are relatively weak.

Figure S1

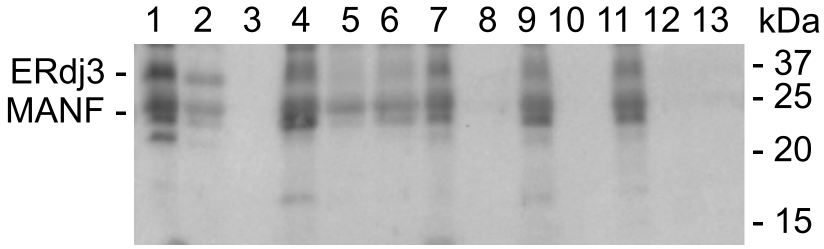

### Figure S2

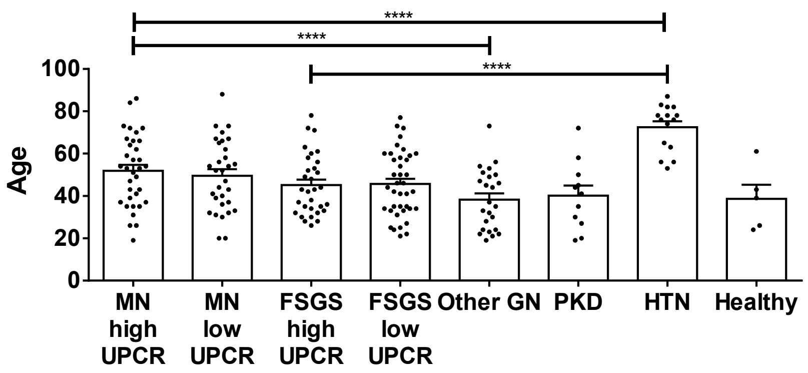

****
****
****
100
80
60
40
20
0
Age
MN high UPCR
MN low UPCR
FSGS high UPCR
FSGS low UPCR
Other GN
PKD
HTN
Healthy

### Figure S3

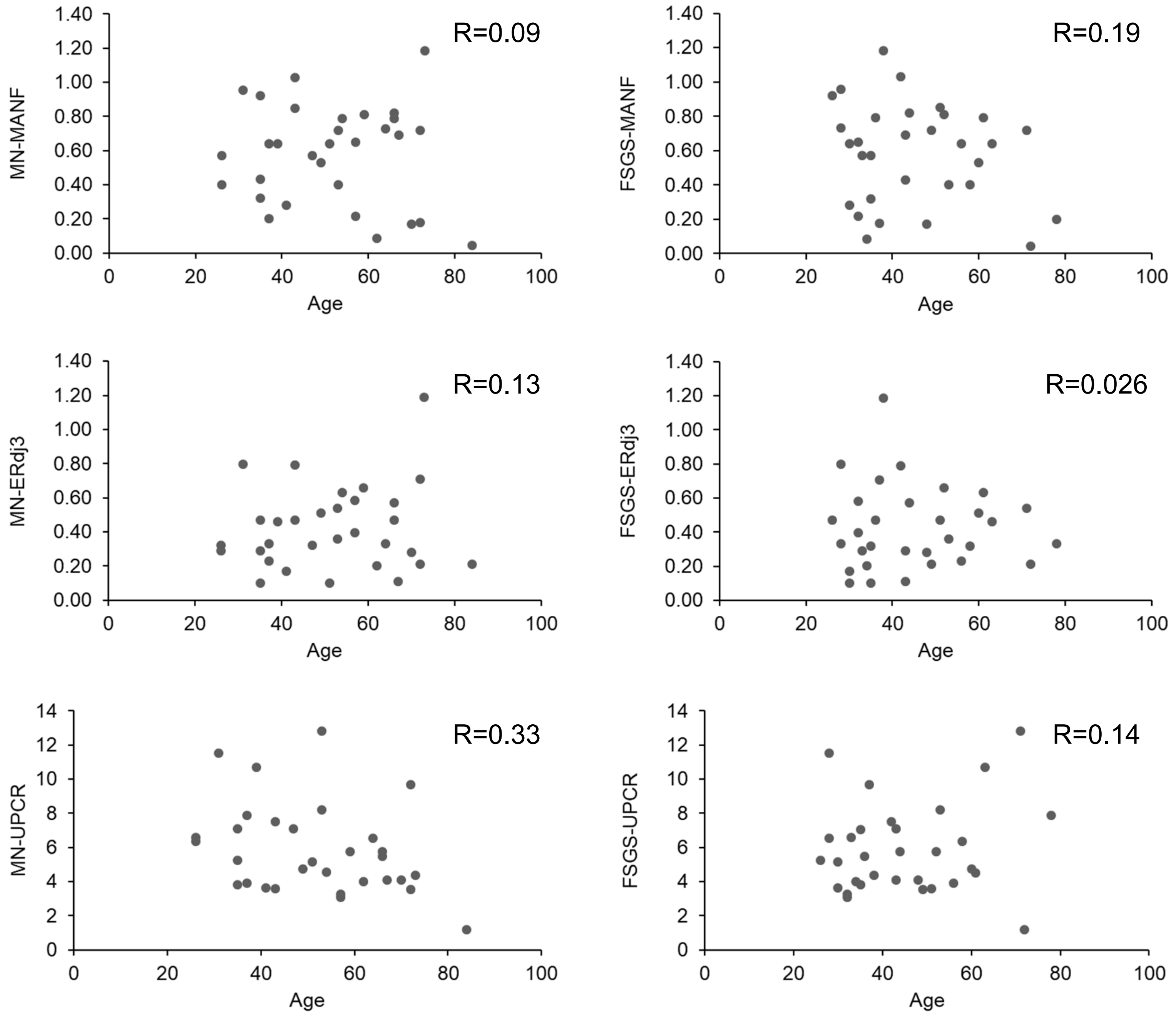
R=0.09
MN-MANF
Age
R=0.19
FSGS-MANF
Age
R=0.13
MN-ERdj3
Age
R=0.026
FSGS-ERdj3
Age
R=0.33
MN-UPCR
Age
R=0.14
FSGS-UPCR
Age
